# Investigate the causality and genetic association between migraine and Parkinson’s disease

**DOI:** 10.1101/2024.01.31.24302057

**Authors:** Ming-Gang Deng, Xiuxiu Zhou, Fang Liu, Kai Wang, Lingli Luo, Min-Jie Zhang, Qianqian Feng, Jiewei Liu

## Abstract

**Background:** The relationship between migraine and Parkinson’s disease (PD) remains controversial. We intend to investigate the causal and genetic association between migraine and PD.

**Methods:** Data related to migraine [any migraine (AM), migraine without aura (MO), and migraine with aura (MA)] and PD were respectively obtained from the latest Genome-wide meta-analysis conducted by the International Headache Genetics Consortium and the International Parkinson’s Disease Genomics Consortium. Univariate Mendelian Randomization (UMR) and multivariate MR (MVMR) were conducted to estimate their bidirectional causality, and global/local genetic correlation and tissue expression analyses were utilized to assess their genetic associations.

**Results:** The UMR presented that AM (OR: 1.016, 95% CI: 0.908-1.135, *p* = 0.785), MO (OR: 0.944, 95% CI: 0.836-1.067, *p* = 0.358) and MA (OR: 1.006, 95% CI: 0.951, 1.064, *p* = 0.846) were unlikely to be associated with PD risk. Similarly, the reverse UMR analyses demonstrated that PD was unrelated to the risks of migraine and its subtypes. These null associations were supported by the MVMR when adjusted for coronary heart disease and hypertension. The global genetic correlation analyses presented that AM (*r*_g_ = -0.061, *p* = 0.127), MA (*r*_g_ = -0.047, *p* = 0.516), MO (*r*_g_ = -0.063, *p* = 0.492) were generally not related to PD, and the local genetic correlation analyses shown they were also uncorrelated in any region. Additionally, the tissue expression analyses found they had no shared tissues.

**Conclusion:** This study suggested no causality or association between migraine and PD from the genetic perspective.

## 1. Introduction

Migraine is a highly prevalent brain disorder characterized by recurrent headache attacks, often accompanied by symptoms such as photophobia, phonophobia, nausea, and vomiting^1^. According to whether the attack was preceded by an aura, migraine could be further classified into migraine without aura (MO) and migraine with aura (MA)^1^. Recent studies have confirmed that migraine is bi-directionally related to the risk of multiple diseases including major depressive disorder^2^, epilepsy^3^, coronary artery disease^4^, etc.

Parkinson’s disease (PD) is a chronic progressive neurodegenerative movement disorder characterized by a profound and selective loss of nigrostriatal dopaminergic neurons^5^. Considering the shared involvement that has been reported among neurotransmitters, the brainstem, and hypothalamic regions in both migraine and PD, many epidemiology studies explored their relationship but have not obtained consent yet. Some studies concluded that migraine was not related to the risk of PD^6-8^, nevertheless, recent studies found people with migraine were more susceptible to suffering from PD^9,10^. Meanwhile, Nunes *et al*. demonstrated that PD patients had a lower lifetime prevalence of headache than controls^11^, and a multi-center case-controlled study found the overall headache and migraine severity reduced after PD onset^12^, which together suggested a potential causal effect of PD on migraine. Therefore, the recent meta-analysis stated that the exact causal relationship between migraine and PD was still ambiguous^13^.

Migraine and PD have both been reported to have substantial genetic backgrounds^14,15^, and the recent genome-wide association studies (GWASs) have identified plenty of genome-wide significant genetic variations related to migraine^16,17^ and PD^18^, which provided an opportunity to investigate their relationship from the genetic perspective. Mendelian Randomization (MR), which uses genetic variants associated with specific exposures as the instrumental variables, could overcome the limitations of conventional observational studies such as reverse causality and residual confusion^19^ to provide a more robust causal estimate of exposures on outcomes because germline genetic variants are randomly inherited from parents to offspring^20^. Therefore, we would like to utilize MR to examine the bidirectional causal relationship between migraine and PD based on the GWAS summary statistics. Genetic correlation and tissue enrichment analyses were also conducted to seek their genetic associations. We hypothesize that migraine and PD were not correlated from the genetic perspective based on current data.

## 2. Methods

### 2.1. Data sources

This study is designed to investigate the bidirectional causal relationship and the genetic association between migraine and PD, an overview of this study design is presented in **Supplementary Fig. 1**. Genetic variants associated with any migraine (AM) were obtained from the latest Genome-wide meta-analysis by Hautakangas *et al*^16^, which included a total of 102,084 cases and 771,257 controls from the International Headache Genetics Consortium (IHGC)-2016^17^, 23andMe, UK Biobank, GeneRISK, and Nord-Trøndelag Health Study. In this meta-analysis, migraine was defined as self-reported or based on the International Classification of Headache Disorders second edition criteria. Due to the data release policy, we retrieved the summary-level statistics with the samples from 23andMe being excluded, and this elimination resulted in a subsample of 589,356 (48,975 cases and 540,381 controls).

Additionally, to evaluate the relationship between PD and migraine subtypes [migraine without aura (MO) and migraine with aura (MA)], genetic data were obtained from the IHGC-2016^17^. MO participants consisted of 8,348 cases and 139,622 controls from 11 studies from the Danish Headache Center, German migraine without aura, etc. MA samples composed of 6,332 cases and 144,883 controls from 12 studies including Danish Headache Center, Dutch Migraine with aura, Nord-Trøndelag Health Study, etc. Detailed information can be found in the original paper^17^.

PD associated genetic variants were obtained from the International Parkinson’s Disease Genomics Consortium^18^, which included 37,688 cases, 18,618 UK Biobank proxy cases, and about 1.4 million controls from 17 cohorts. As GWAS results of several datasets (such as 23andMe) were not publicly available, genome-wide associations of 33,674 cases and 449,056 controls were used in this study.

### 2.2. Mendelian Randomization

To ensure the causal estimates from the MR are valid, the genetic variants should satisfy three critical assumptions: (1) the relevance assumption indicates the genetic variants are strongly associated with the exposure, (2) the independence assumption implies genetic variants are not associated with any potential confounder of the exposure-outcome association and (3) the exclusion restriction assumption refers that variants do not affect outcome independently of exposure^21^. For the bidirectional MR causal estimates, the analyses were performed in two directions with genetic variants with migraine (1) as exposure: to evaluate whether individuals suffered from migraine were more likely to develop PD, and (2) as outcome: to assess whether people with PD were less likely to report migraine.

The procedures of instrument selection in our study were in line with the aforementioned three critical assumptions. For the relevance assumption, SNPs were initially screened at genome-wide significance (*P* < 5×10^−8^), and if no SNP were discovered, this threshold was loosened to *P* < 1×10^−5^ as previous studies did^2,22^. To avoid the bias triggered by weak instruments, the F statistic was calculated and SNPs with an F statistic lower than 10 were considered as weak instruments and excluded^23,24^. For the exclusion restriction assumption, the retained SNPs were clumped for linkage disequilibrium (LD) with the European samples from the 1000 Genomes Project^25^ as the reference panel using the PLINK tool (version 1.9)^26^. The clumping R^2^ cut-off was set as 0.001 within a window of 10 Mb. If an obvious LD effect was detected, only the SNP with the lowest *p*-value would be kept. Additionally, the modified Cochran’s Q test^27^ was performed to identify outlier pleiotropic SNPs as the Q statistic much larger than N_SNP_-1 suggests violation of independence or exclusion restriction assumption^28^, and these outlier pleiotropic SNPs would be excluded. As for the independence assumption, we used the PhenoScanner V2 tool^29^ to assess whether the selected SNPs were related to the confounders or risk factors in the relationship between migraine and PD. If obvious confounders were identified, they would be further adjusted in the multivariate MR (MVMR) analyses, which is an extension of MR that can detect the causal effects of multiple risk factors jointly^30^. The inverse variance weighted (IVW) method was adopted as the primary method to infer the causal relationship between migraine and PD, which used a meta-analysis approach that combined the Wald estimate for each SNP to obtain an overall estimate and was the most efficient method when average pleiotropic effect did not exist. Additionally, three well-established and horizontal pleiotropy robust methods including MR-Egger, weighted median, and MR-PRESSO (Pleiotropy RESidual Sum and Outlier) were supplemented to validate the IVW estimates. Sensitive analyses by the MR-Egger regression intercept and leave-one-out analyses were respectively conducted to check whether directional horizontal pleiotropy or a single SNP was driving the results of MR analyses.

The MR analyses were performed in R software (version 4.3.1) with *TwoSampleMR* (version 0.5.7), *MendelianRandomization* (version 0.8.0), *MRPRESSO* (version 1.0), and *RadialMR* (version 1.1) packages.

### 2.3. Genetic association analyses

#### 2.3.1. Global Genetic Correlation

The LD score regression^31^, which could evaluate the genetic heritability correlation cross traits based on GWAS summary statistics, was adopted to evaluate the genetic correlation (*r*_g_) between migraines and PD, with European ancestry samples from the 1000 Genomes Project being the reference panel^25^. This procedure was performed in R software (version 4.3.1) with the *ldscr* (version 0.1.0) package.

### 2.3.2. Local Genetic Correlation

To investigate whether migraine and PD were locally correlated at a defined genomic region, *ρ*-HESS (Heritability Estimation from Summary Statistics)^32,33^, which was a software package for estimating and visualizing local SNP-heritability and genetic correlation from GWAS summary association data, was utilized. In this algorithm, the entire genome was partitioned into 1703 independent regions across all chromosomes (sex chromosomes excluded) based on the European population LD patterns. The local genetic correlation between migraine and PD was quantified in particular regions, and the Bonferroni corrected *P*-value of *P* < 0.05/1703 was set as the significance threshold.

#### 2.3.3. Tissue expression analyses

To explore whether the GWAS data related to migraine and PD were enriched in some specific and same tissues, we implemented the MAGMA (Multi-marker Analysis of GenoMic Annotation)^34^ in FUMA^35^ (https://fuma.ctglab.nl/) to perform the tissue expression analyses, which was based on the expression data from 54 tissues of GTEx V8^36^. The maximum *p*-value of lead SNPs was set as *P* < 5×10^−8^, and if no SNP met this criterion, we loosened it to *P* < 1×10^−5^. The upstream and downstream windows size of genes to assign SNPs were set to 35 and 10kb^37^, respectively. The other parameters and settings were used default. Bonferroni correction was applied and a *P*-value threshold of *P* < 0.05/54 was set to obtain significant findings.

## 3. Results

### 3.1. Mendelian Randomization

#### 3.1.1. Univariate Mendelian Randomization

The selected instrumental variants based on prior criteria were presented in **Supplementary Table 1-2**. The total proportion of variance explained by their corresponding data sets ranged from 3.986% to 21.772%.

The results of MR causal inference between migraine and PD are illustrated in **Table 1**. The IVW method presented that genetically predicted liability to AM (OR: 1.016, 95% CI: 0.908-1.135, *P* = 0.785), MO (OR: 0.944, 95% CI: 0.836-1.067, *P* = 0.358) and MA (OR: 1.006, 95% CI: 0.951-1.064, *P* = 0.846) were unlikely to be associated with PD risk. Regarding the reverse association, the IVW method also demonstrated that PD was not related to the risk of AM (OR: 1.003, 95% CI: 0.975-1.032, *P* = 0.817), MO (OR: 0.956, 95% CI: 0.893-1.025, *P* = 0.207) and MA (OR: 1.032, 95% CI: 0.955-1.115, *P* = 0.430). These null associations were generally supported by the supplemented analyses by MR-Egger, weighted median, and MR-PRESSO methods. The scatter plot of SNP effects on migraine versus PD, and PD versus migraine was presented in **Supplementary Fig. 2**. Sensitive analyses by the MR-Egger regression intercept showed there was no directional horizontal pleiotropy affecting the overall causal estimates (**Table 1**) and leave-one-out analyses implied the overall effect was not driven by a single SNP (**Fig. 2**).

**Table 1.** Univariate Mendelian Randomization to infer the bidirectional relationship between migraine and Parkinson’s disease.

| Exposure | Outcome | N<sub>SNP</sub> | Methods | OR (95% CI) | *p*-value | Cochran's Q | *p*-value for heterogeneity<sup>†</sup><br>or pleiotropy<sup>‡</sup> |
| --- | --- | --- | --- | --- | --- | --- | --- |
| AM | PD | 32 | IVW | 1.016 (0.908, 1.135) | 0.785 | 35.320 | 0.271 |
|  |  |  | MR Egger | 0.828 (0.602, 1.139) | 0.256 |  | 0.192 |
|  |  |  | Weighted median | 0.967 (0.828, 1.130) | 0.676 |  |  |
|  |  |  | MR-PRESSO | 1.016 (0.908, 1.135) | 0.787 |  |  |
| MO | PD | 5 | IVW | 0.944 (0.836, 1.067) | 0.358 | 1.302 | 0.861 |
|  |  |  | MR Egger | 0.571 (0.189, 1.725) | 0.394 |  | 0.435 |
|  |  |  | Weighted median | 0.962 (0.828, 1.118) | 0.614 |  |  |
|  |  |  | MR-PRESSO | 0.944 (0.881, 1.012) | 0.182 |  |  |
| MA | PD | 34 | IVW | 1.006 (0.951, 1.064) | 0.846 | 31.330 | 0.550 |
|  |  |  | MR Egger | 1.013 (0.887, 1.156) | 0.853 |  | 0.858 |
|  |  |  | Weighted median | 0.987 (0.910, 1.071) | 0.755 |  |  |
|  |  |  | MR-PRESSO | 1.020 (0.959, 1.085) | 0.529 |  |  |
| PD | AM | 19 | IVW | 1.003 (0.975, 1.032) | 0.817 | 18.355 | 0.433 |
|  |  |  | MR Egger | 0.932 (0.851, 1.019) | 0.142 |  | 0.107 |
|  |  |  | Weighted median | 0.990 (0.950, 1.031) | 0.625 |  |  |
|  |  |  | MR-PRESSO | 1.003 (0.975, 1.032) | 0.819 |  |  |
| PD | MO | 19 | IVW | 0.956 (0.893, 1.025) | 0.207 | 14.552 | 0.692 |
|  |  |  | MR Egger | 0.829 (0.692, 0.994) | 0.043 |  | 0.113 |
|  |  |  | Weighted median | 0.932 (0.845, 1.029) | 0.163 |  |  |
|  |  |  | MR-PRESSO | 0.956 (0.899, 1.018) | 0.177 |  |  |
| PD | MA | 18 | IVW | 1.032 (0.955, 1.115) | 0.430 | 6.761 | 0.986 |
|  |  |  | MR Egger | 0.921 (0.764, 1.109) | 0.398 |  | 0.206 |
|  |  |  | Weighted median | 1.038 (0.932, 1.156) | 0.494 |  |  |
|  |  |  | MR-PRESSO | 1.032 (0.982, 1.083) | 0.228 |  |  |

<sup>†</sup> *p*-value for heterogeneity based on Cochran's Q statistic.
<sup>‡</sup> *p*-value or pleiotropy based on MR-Egger regression intercept.
Abbreviations: AM: any migraine; CI: confidence interval; MA: migraine with aura; MO: migraine without aura; IVW: inverse variance weighted; OR: odds ratio; PRESSO: Pleiotropy RESidual Sum and Outlier; SNP: Single-nucleotide polymorphism.

**Fig 1.**
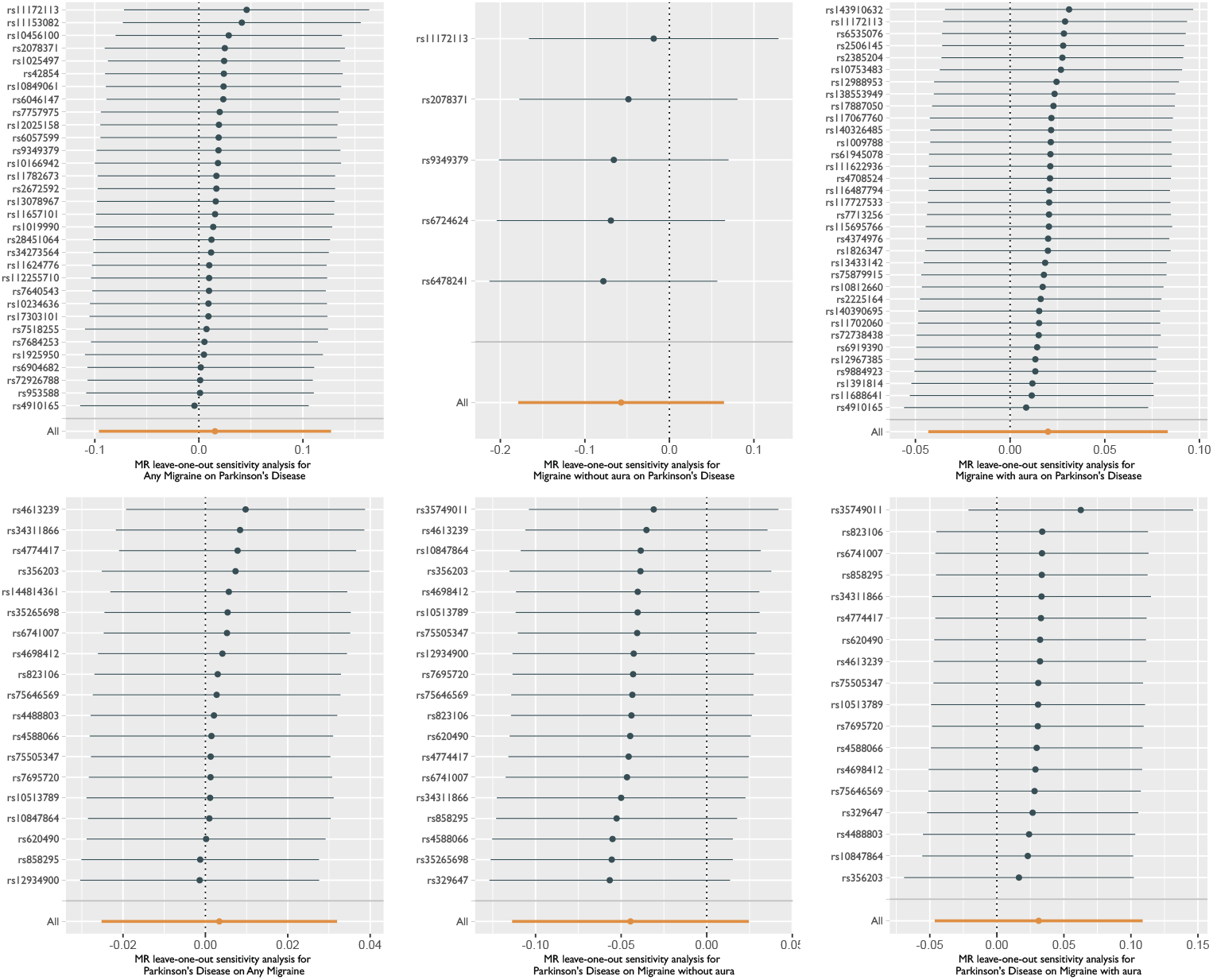
MR leave-one-out analyses of migraine on Parkinson’s disease (above) and Parkinson’s disease on migraine (below).

**Fig 2.**
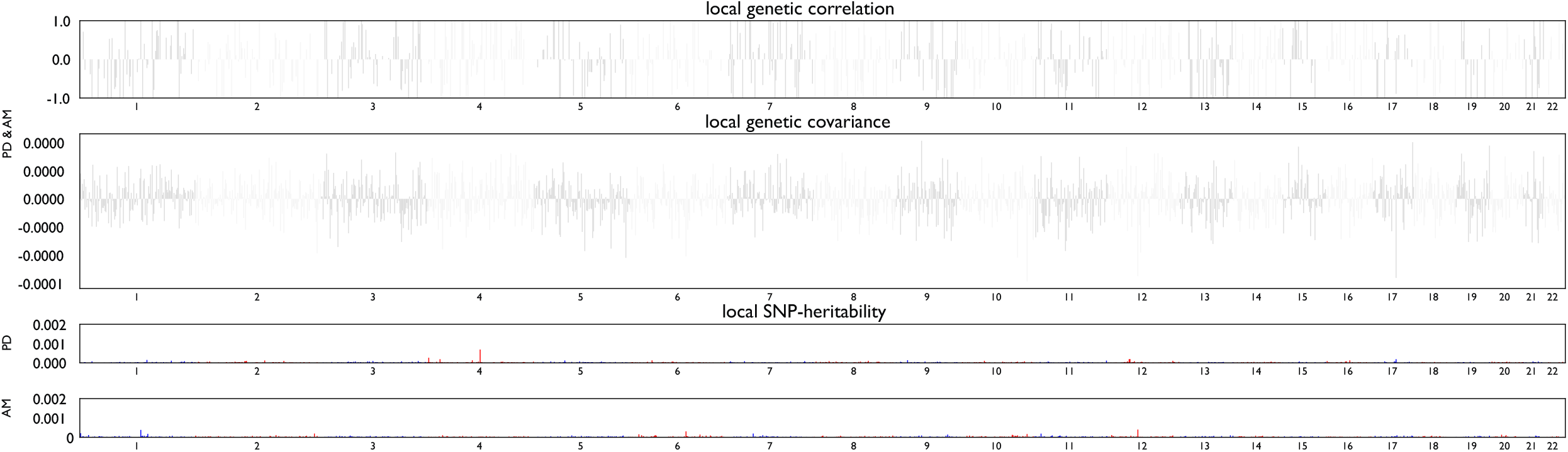
Local genetic correlation and genetic covariance between any migraine and Parkinson’s disease, and also local SNP heritability estimation for migraine and Parkinson’s disease respectively.

#### 3.1.2. Multivariate Mendelian Randomization

As shown in **Supplementary Table S1-S2**, genetic variants associated with migraine were meanwhile related to coronary heart disease and hypertension, which have been demonstrated as risk factors of PD^38,39^, while no obvious confounding phenotypes were identified in genetic variants related to PD. Therefore, we performed MVMR analyses to control the influence of coronary heart disease and hypertension and the result is presented in **Table 2**.

**Table 2.**
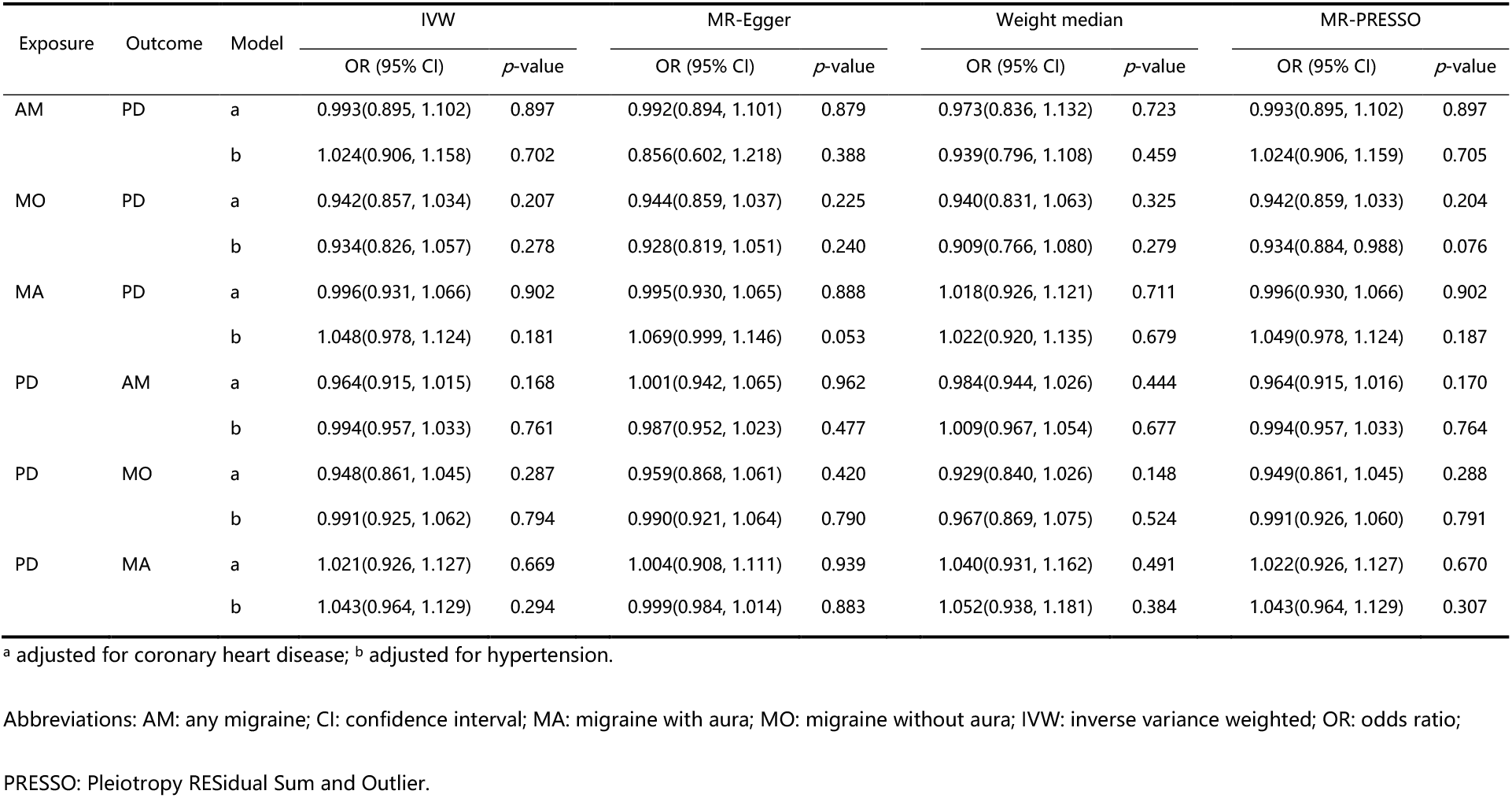
Multivariate Mendelian Randomization to investigate the bidirectional causal relationship between migraine and Parkinson’s disease.

After adjustment for coronary heart disease, the IVW method indicates AM (OR: 0.993, 95% CI: 0.895-1.102, *P* = 0.897), MO (OR: 0.942, 95% CI: 0.857-1.034, *P* = 0.207), and MA (OR: 0.996, 95% CI: 0.931-1.066, *P* = 0.902) were not associated with PD risk, and reversely PD was also not correlated with migraine risks. This bidirectional null association was supported by the results found by MR-Egger, weighted median, and MR-PRESSO methods. Similarly, migraine was not associated with PD risk and vice-versa when controlled for the confounding effect of hypertension.

#### 3.1.3. Assessment of assumptions

Firstly, for the relevance assumption, instrumental variables related to migraine and PD were selected from the recent GWASs with large sample sizes, and SNPs with an F statistic of less than 10 were excluded, thus making it unlikely to induce weak instrument bias. Secondly, for the independence assumption, the selected SNPs were identified to be related to coronary heart disease and hypertension, which were the confounding factors in the association between migraine and PD. To avoid this bias, we performed the MVMR to control the confounding effect, and the association between migraine and PD was still robust. Thirdly, for the exclusion restriction assumption, we clumped SNPs at a restricted standard to make the SNPs independent of each other, and modified Q tests were conducted to identify the outlier pleiotropic SNPs. Besides, the MR-Egger regression intercept term indicated there was no significant horizontal pleiotropy among the instruments.

### 3.2. Genetic association analyses

Although no obvious causality was found between migraine and PD, we further attempted to investigate whether they were genetically correlated via global/local genetic correlation and tissue expression analyses.

#### 3.2.1. Global Genetic Correlation

We used LDSC to investigate the global genetic correlation between PD and migraine based on their corresponding GWAS summary statistics. The LD score regression showed that AM (*r*_g_ = -0.061, *P* = 0.127), MA (*r*_g_ = -0.047, *P* = 0.516), MO (*r*_g_ = -0.063, *P* = 0.492) were generally not related to PD based on GWASs results.

#### 3.2.2. Local Genetic Correlation

Although we didn’t observe a significant genetic correlation between PD and migraine (including AM, MA, and MO), we then tested whether PD had a genetic correlation with migraine locally. We used *ρ*-HESS to investigate the local genetic correlation between PD and migraine. The results of local genetic correlation analyses between AM and PD are presented in **Fig. 2** and **Supplementary Table S3**. After correcting for multiple testing (*P* <0.05/1703), no significant local genetic correlation between AM and PD was observed. As for the migraine subtypes, we discovered MO and PD (**Supplementary Fig. 3 and Supplementary Table S4**) or MA and PD (**Supplementary Fig. 4 and Supplementary Table S5**) were also not locally correlated in any specific genomic region.

#### 3.2.3. Tissue expression analyses

The tissue expression analyses showed GWAS associations related to AM were significantly enriched in colon sigmoid tissue, while GWAS data related to PD were significantly enriched in 6 brain tissues, with the most significant enrichment in Frontal Cortex BA9 (**Fig. 3**). However, genetic data related to MO and MA were not enriched in any GTEx v8 tissue (**Supplementary Fig. S5**). Based on the above results, we discovered that the expression of migraine and PD did not share any same tissue.

**Fig 3.**
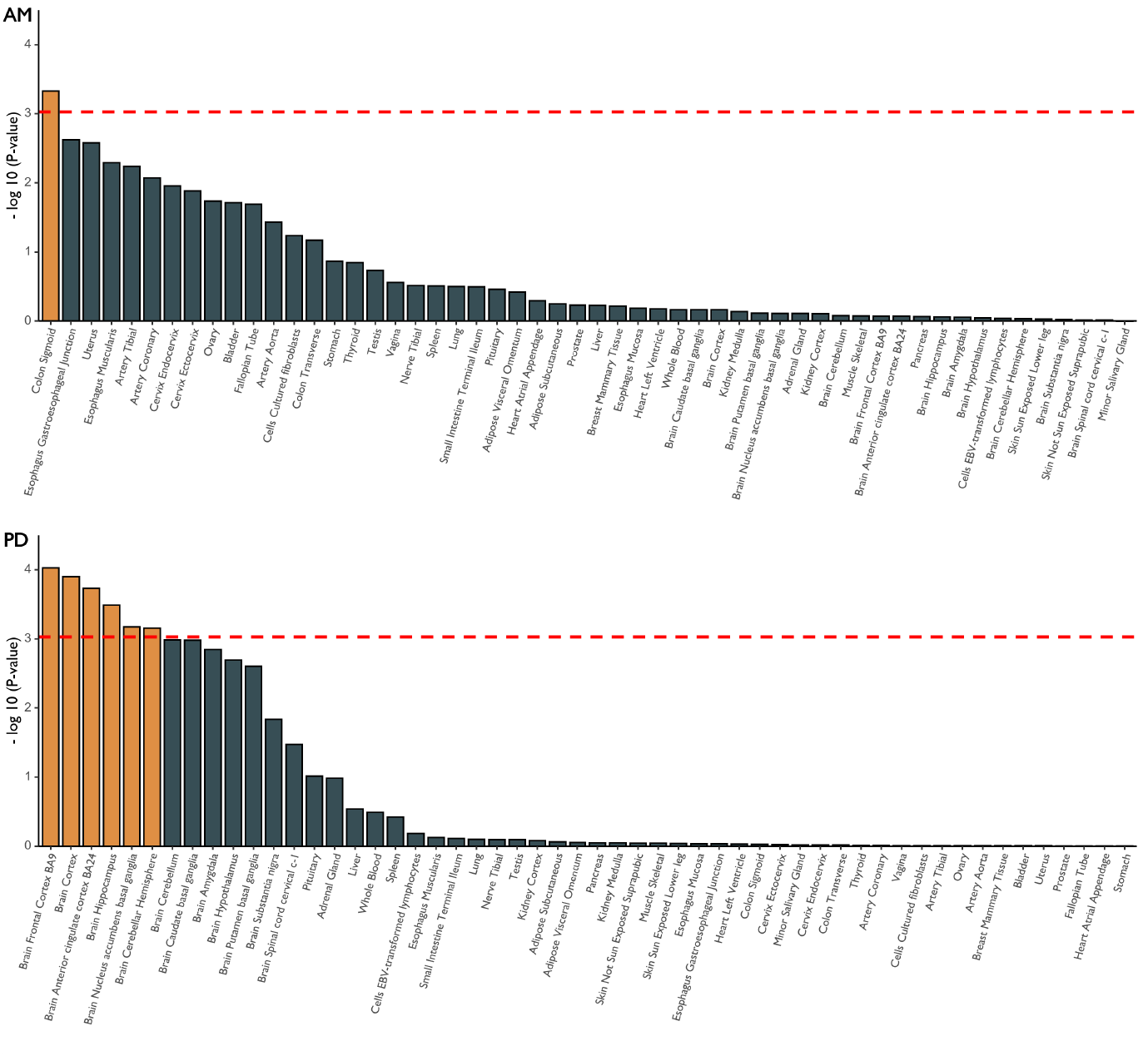
Tissue-type enrichments of AM and PD GWAS associations. The red dashed line is the Bonferroni corrected significant level. Tissues that showed significant enrichment (corrected *P* < 0.05) are shown in yellow. AM = Any migraine; PD = Parkinson’s disease.

## 4. Discussion

Using the latest and largest GWAS summary data related to migraine and PD, we assessed their bidirectional causality via the univariate and multivariate MR analyses but failed to support their causal relationship. We further investigated their genetic association through the global/local genetic correlation and tissue expression analyses, and the results indicated migraine and PD were unlikely to be genetically correlated.

Previous observational studies have tried to investigate the relationship between migraines and PD^6-13^, but the sample size was relatively limited and the findings were contradictory. The possible mechanism to support the relationship between migraine and PD is mainly reflected in dopaminergic pathways. It was suggested that people who suffered from migraine had chronic dopaminergic hypofunction and dopamine receptor hypersensitivity^10,40^, and the degeneration of dopamine neurons in dopaminergic pathways is the key hallmark of PD^15,41^. In this study, we aimed at exploring the causality and genetic relationship between migraine and PD based on GWAS results. Our MR and genetic correlation analyses didn’t support a significant causality and genetic relationship between migraine and PD. A possible explanation is that over 200 genes that participate in broad dopaminergic pathways^42^, migraine, and PD may be affected by different genes among these genes to dysregulate dopaminergic pathways.

Our study has several strengths that are worth pointing out. First of all, our GWAS data related to migraine and PD both came from the recent large-scale surveys, the sample sizes of which were much larger than previous observational studies. In addition, the associations between PD and migraine subtypes were investigated in this study, which was neglected by previous research. Furthermore, the MR and genetic association analyses could avoid reverse causality and confounding bias from conventional epidemiology studies.

Despite the above advantages, the findings from our study should be interpreted within the context of several limitations. For example, the GWAS sample size of PD and migraine needs further extension, e.g., the 23andMe samples are not included in publicly available GWAS summary statistics due to the data release policy of 23andMe^16,18^. Besides, GWASs could only capture common genetic variations that related to diseases, variations like copy number variations^43,44^ and rare mutations^45,46^ were not included, which are also important components of the genetic architecture of diseases. Lastly, the genetic data we used are largely Europeans, further GWASs of PD and migraine performed in non-European populations are warranted to confirm our results in this study.

To sum up, we performed a series of analyses including MR, genetic correlation, and tissue enrichment analyses to investigate the causal and genetic relationship between migraine and PD based on their GWAS datasets. Our result didn’t find clues for the relationship between migraine and PD by current GWAS results, however, this conclusion needs further confirmed in larger sample GWASs and different ethnic groups.

## Data Availability

GWAS summary statistics for migraine and subtypes were available at the International Headache Genetics Consortium (http://www.headachegenetics.org/), and Parkinson's disease associated data could be obtained from the International Parkinson's Disease Genomics Consortium (https://www.pdgenetics.org/).

## 5. Contributors

**J.L**. designed the study and edited the manuscript. **M.-G.D**. designed the study, performed the statistical analysis, and drafted the manuscript. **X.Z**. drafted and edited the manuscript. **F.L**., **K.W**., **L.L**., **M.-J.Z**., **and Q.F**. reviewed and edited the manuscript.

## 7. Acknowledgments

We acknowledge the participants from the International Headache Genetics Consortium.

## 8. Data Sharing Statement

GWAS summary statistics for migraine and subtypes were available at International Headache Genetics Consortium (http://www.headachegenetics.org/), and Parkinson’s disease associated data could be obtained from the International Parkinson’s Disease Genomics Consortium (https://www.pdgenetics.org/).

## 10. Ethics approval

The data sets used were all public available, and the ethical approvals were obtained in the original papers.

## 11. Consent to participate/publish

Not applicable.

