## Supplementary Figures for "Investigate the causality and genetic association between migraine and Parkinson’s disease"

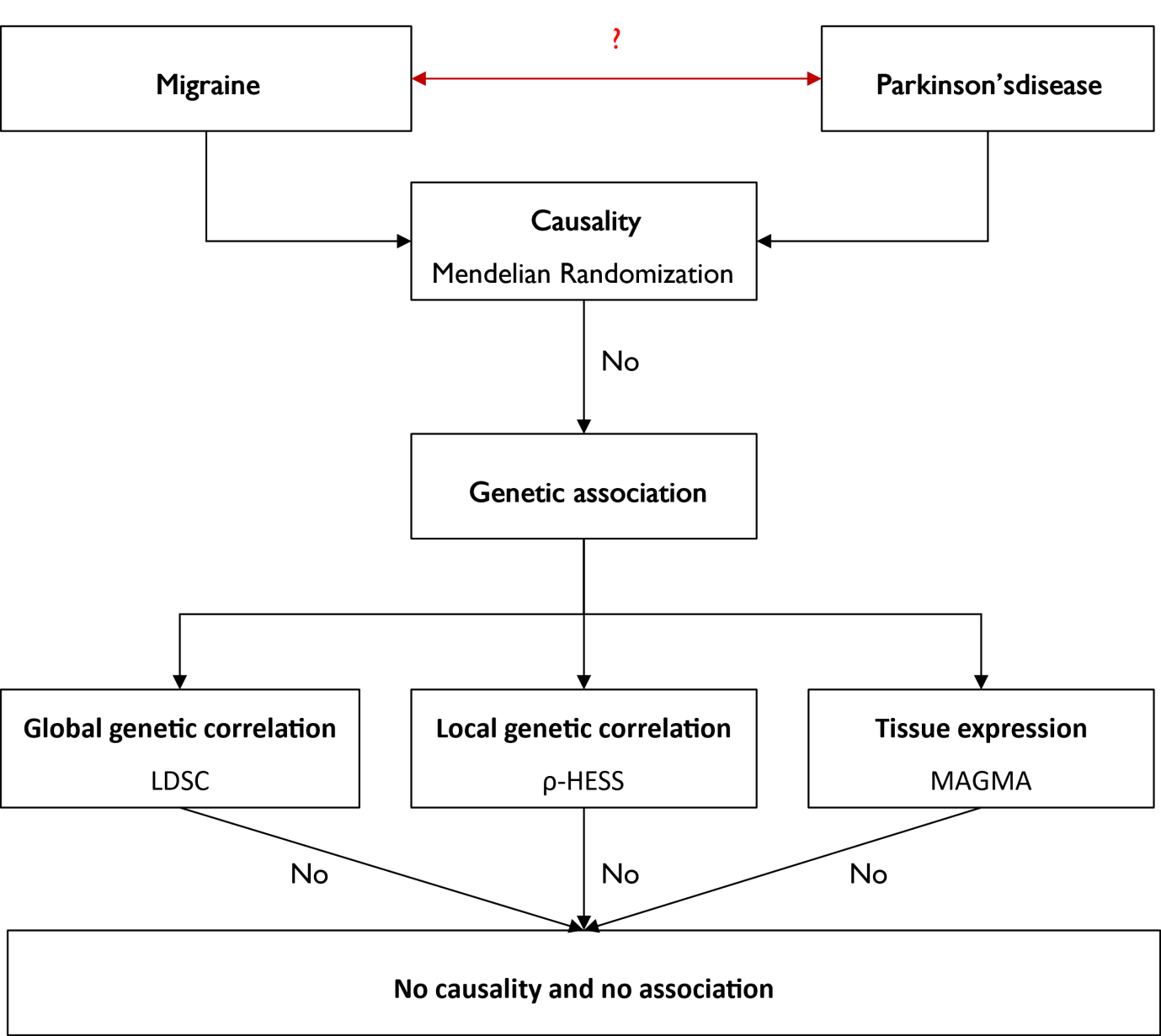

**Supplementary Fig. 1** The diagram of the study design.

Abbreviations: LDSC, linkage disequilibrium score regression; HESS, heritability estimation from summary statistics; MAGMA, Multi-marker Analysis of GenoMic Annotation.

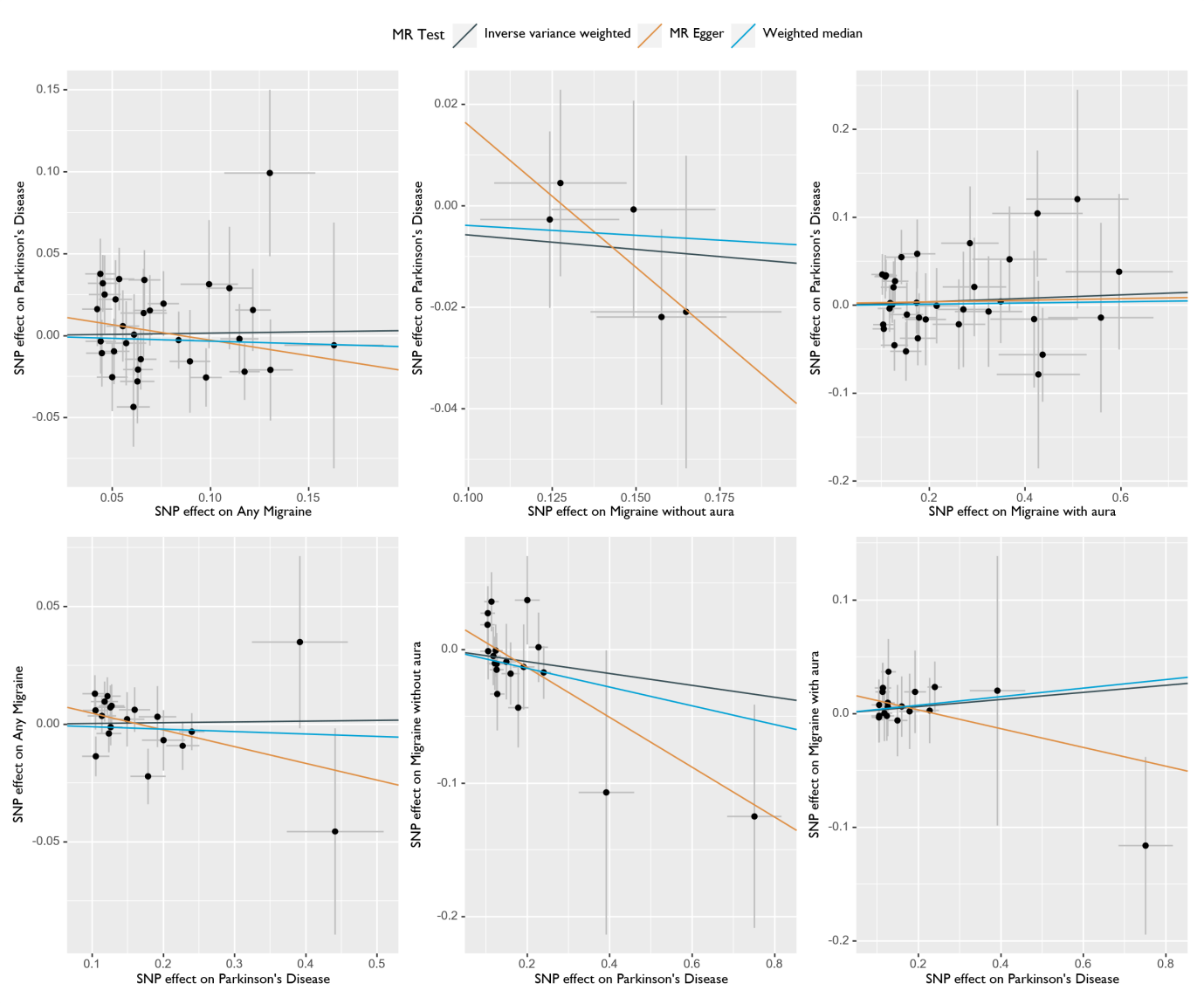

**Supplementary Fig 2.** Scatter plots of SNP effects on migraine versus Parkinson’s disease (above), and Parkinson’s disease versus migraine (below), with the slope of each line corresponding to the estimated MR effect per method.

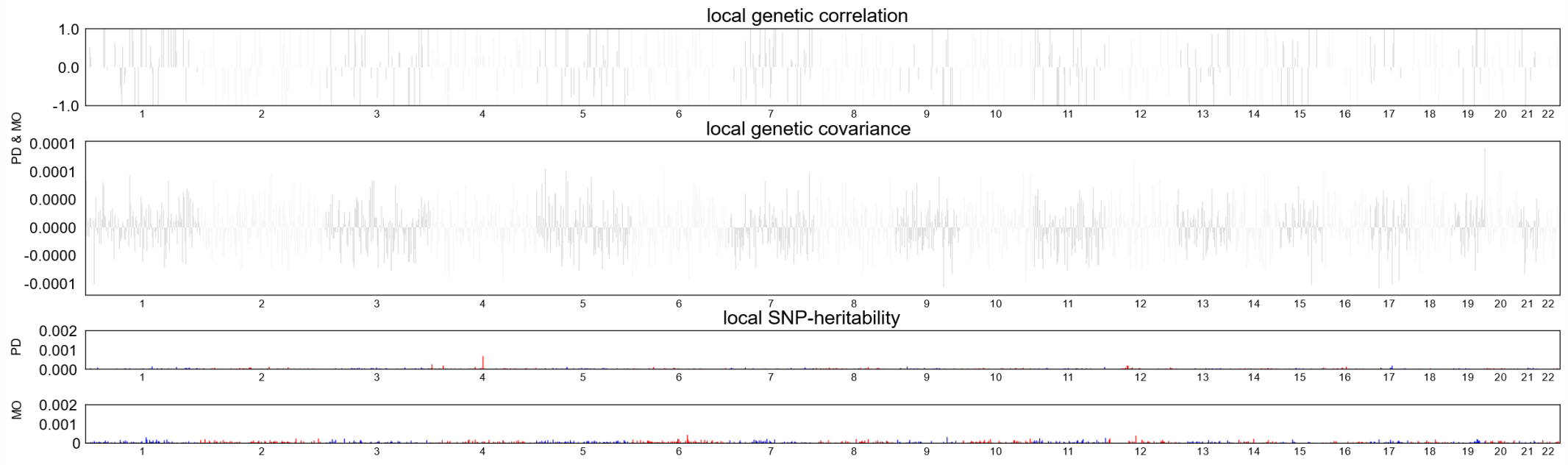

**Supplementary Fig 3.** Local genetic correlation, genetic covariance, and SNP heritability between MO and PD.

Abbreviations: MO, migraine without aura; PD, Parkinson’s disease.

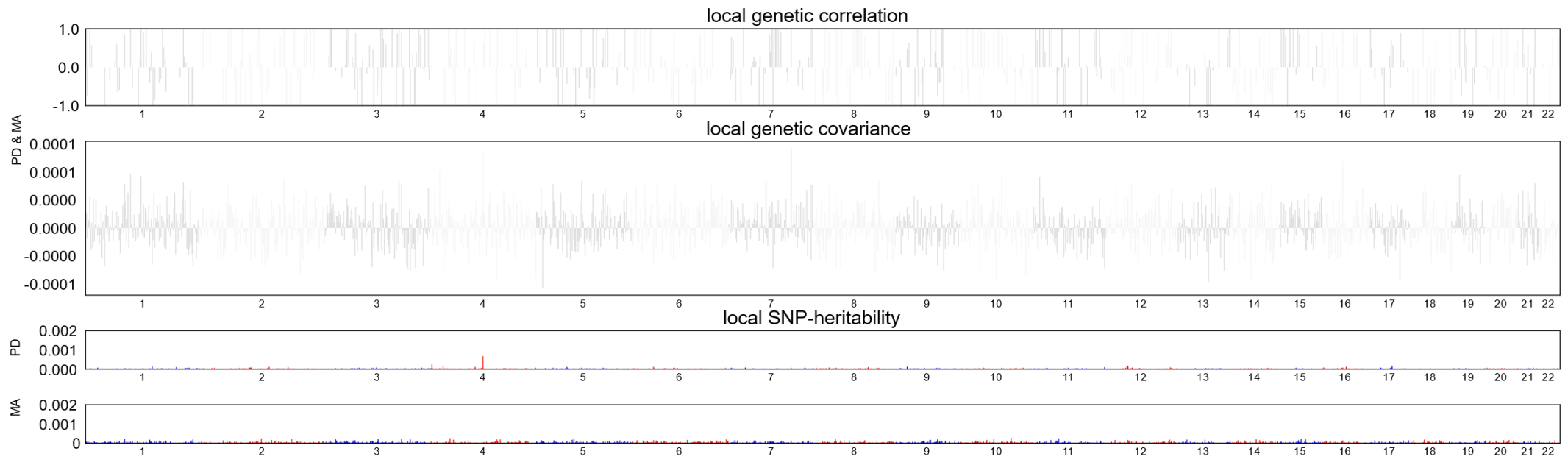

**Supplementary Fig 4.** Local genetic correlation, genetic covariance, and SNP heritability between MA and PD.

Abbreviations: MA, migraine with aura; PD, Parkinson’s disease.

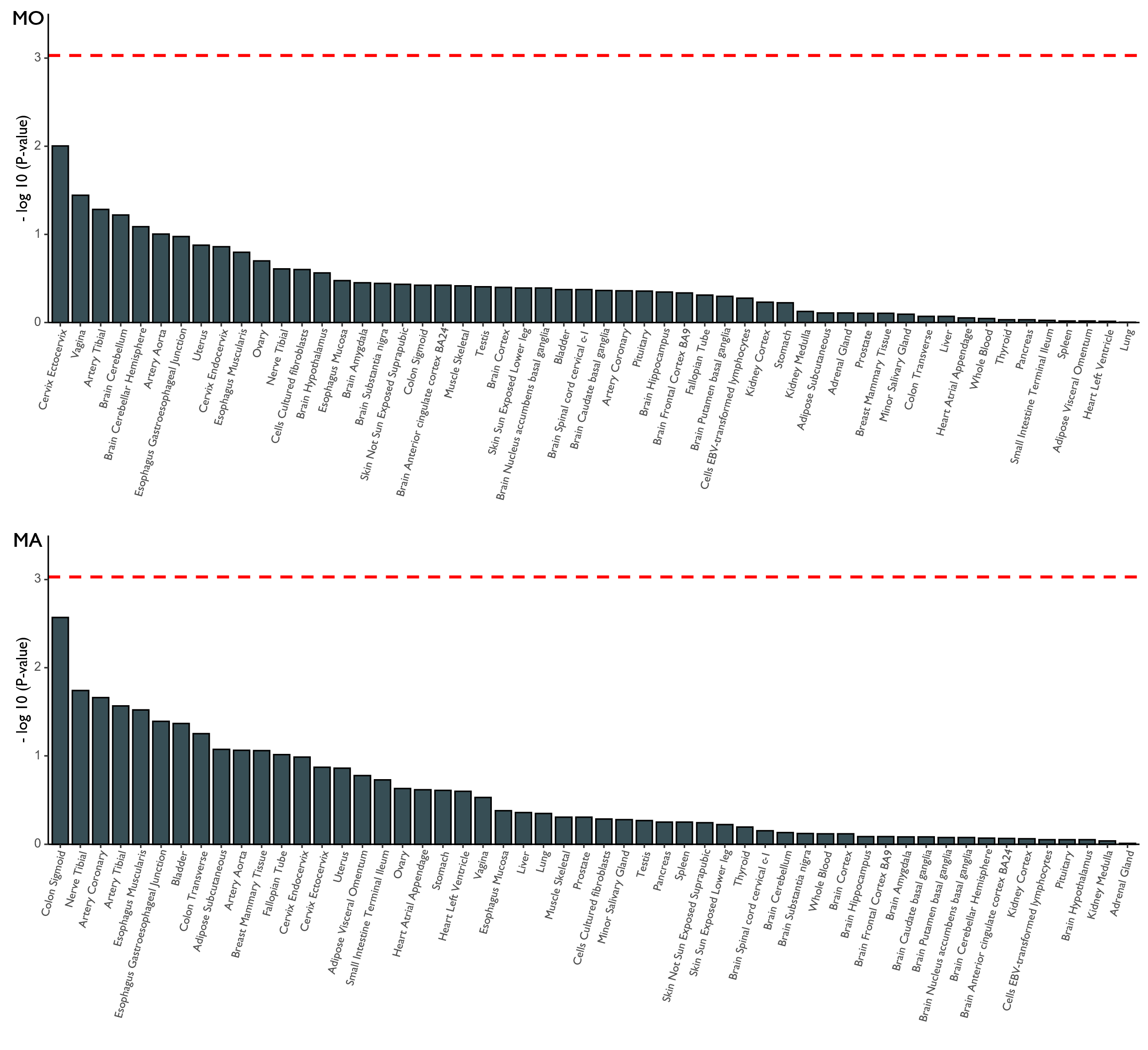

**Supplementary Fig 5.** Tissue-type enrichments of MO and MA GWAS associations. The red dashed line is the Bonferroni corrected significant level. Tissues that showed significant enrichment (corrected *p* < 0.05) are shown in yellow.

Abbreviations: MA: migraine with aura; MO, migraine without aura.
